# Prediction of COVID-19 mortality among hospitalized patients in Sudan

**DOI:** 10.1101/2021.03.09.21253179

**Authors:** Ghada Omer Hamad Abd El-Raheem, Maysoun Ahmed Awad Yousif, Doaa Salih Ibrahim Mohamed

## Abstract

**Background:** COVID-19 was primarily reported in China. The mortality rate across countries had ranged from 1% up to more than 10% and it is underestimated in some countries. Advanced age is the most frequently reported factor associated to mortality. Other factors were the presence of comorbidities such as diabetes mellitus, hypertension and obesity. Several models for mortality prediction had been developed to assist in improving the prognosis. The aim of our study was to assess the factors related to mortality among COVID-19 patients and develop a prediction model based on these factors.

**Methods:** A retrospective cohort study assessed the factors related to the mortality among COVID-19 patients who attended Imperial Hospital isolation centre on November-December, 2020, Khartoum, Sudan. Statistical tests performed were chi-square test, odds ratio and regression to develop the prediction model. Tests were considered statistically significant when *p* < 0.05.

**Results:** 105 patients were studied. 29% of the patients were deceased, while, 71% were discharged alive. A statistically significant association was found between the age and severity with regards to mortality rate (*p*=0.034, 0.018 respectively). The model equation for mortality prediction: Mortality = −14.724+ (1.387* Age) + (−0.323* Gender) + (1.814* Admission) + (0.193* Ischemic Heart Disease) + (−0.369* Fever) + (1.595* Cough) + (1.953* Complications) + (0.149* Duration of hospitalization) + (0.999* Enoxaparin dose).

**Conclusions:** Age, admission ward, cough and enoxaparin dose were statistically significant predictors for COVID-19 mortality (*p*= 0.014, 0.011, 0.015, 0.006 respectively).

## Background

COVID-19 infection is considered a pandemic across the whole world and has been declared as a global public health emergency situation by the world health organization [1, 2]. COVID-19 was primarily reported in China [3]. The mortality rate across countries had ranged from 1% up to more than 10% [4] and it is underestimated in some countries [5]. Advanced age is the most frequently reported factor associated to mortality [4, 6]. Hence focus was applied for older population [6]. Other factors were the presence of comorbidities such as diabetes mellitus (DM), hypertension (HTN) and obesity [2]. COVID-19 infection can lead to multi thrombotic events [7], coagulation abnormality was also proposed to be associated with mortality of COVID-19 patients [8]. Climate factors were associated with the mortality rates of COVID-19 infection as well, such as humidity and temperature especially in African countries [9]. As well as, severe air pollution [10]. Patients with cardiovascular co-morbid conditions were considered vulnerable as well [11]. Several models for mortality prediction had been developed to assist in improving the prognosis [12, 13]. in the other hand, epidemiological models for forecasting the excess mortality and for forecasting the pandemic itself were developed [14, 15]. Many mortality scores were developed to predict COVID-19 outcomes [3, 16, 17]. United nations, OCHA reported that the average COVID-19 daily cases in Sudan had reached 300 cases in December 2020, with Khartoum State accounting for 78% of the reported cases [18]. The aim of our study was to assess the factors related to mortality among COVID-19 patients and develop a prediction model based on these factors.

## Methods

A retrospective cohort study was conducted to assess the factors related to the mortality among COVID-19 patients who attended Imperial Hospital-Isolation centre on November-December, 2020, Khartoum State, Sudan. Imperial Hospital is a 60 bed hospital. It contains three isolation wards. General isolation wards were 2 with 10 beds capacity each, while, the isolation ICU had 5 beds capacity. The sample of 105 files of adult COVID-19 patients was collected randomly from the hospital registry. Patients who attended in November 2020 and December 2020 were involved in the study. The characteristics of the patients and the mortality related risk factors were recorded. Statistical package for social sciences (SPSS version 23) was used to describe and analyse the data. Statistical analysis tests performed were chi-square tests to determine association among variables, odds ratio and regression to develop the prediction model. Tests were considered statistically significant when *p* < 0.05.

## Results

### Mortality status among patients at Imperial Hospital

105 patients were involved in the study. 29% of the patients were deceased, while, 71% were discharged alive.

### Demographic characteristics of COVID-19 patients based on the mortality status

105 patients were involved in the study. Age was assessed with regards to mortality status. None of the patients who aged ≤ 45 years had died. While, (21.1%) of the patients who aged between 46 and 60 years were deceased. In the other hand, the percent of the deceased patients at the age above 60 years was (33.3%). There was a statistically significant association (*p*= 0.034) between mortality rate and the age of the patients.

Regarding gender and comorbid conditions of the patients, there was no statistically significant association between these characteristics and mortality (*p*> 0.05).

Of the patients who were admitted to general isolation wards, 23.5% were deceased. While, among the patients who were admitted to the isolation ICU, 50% were deceased. A statistically significant association (*p*=0.018) was found between the hospital admission and mortality. Table 1 below illustrated the association between the demographic characteristics of the patients and mortality.

**Table 1:** Demographic characteristics of COVI-19 patients based on the mortality status (n=105)

| Characteristics | Mortality |  |  |  | Total | % | Chi<sup>2</sup> | p-value |
| --- | --- | --- | --- | --- | --- | --- | --- | --- |
|  | Deceased | % | Alive | % |  |  |  |  |
| **Age** |  |  |  |  |  |  | 6.784* | 0.034 |
| 30- 45 Years | 0 | 0.0 | 8 | 100.0 | 8 | 7.6 |  |  |
| 46- 60 Years | 4 | 21.1 | 15 | 78.9 | 19 | 18.1 |  |  |
| >60 Years | 26 | 33.3 | 52 | 66.7 | 78 | 74.3 |  |  |
| **Total** | **30** | **28.6** | **75** | **71.4** | **105** | **100.0** |  |  |
| **Gender** |  |  |  |  |  |  | 0.261 | 0.609 |
| Male | 20 | 30.3 | 46 | 69.7 | 66 | 62.9 |  |  |
| Female | 10 | 25.6 | 29 | 74.4 | 39 | 37.1 |  |  |
| **Total** | **30** | **28.6** | **75** | **71.4** | **105** | **100.0** |  |  |
| **Comorbid conditions** |  |  |  |  |  |  | 0.718 | 0.397 |
| Yes | 24 | 30.8 | 54 | 69.2 | 78 | 74.3 |  |  |
| No | 6 | 22.2 | 21 | 77.8 | 27 | 25.7 |  |  |
| **Total** | **30** | **28.6** | **75** | **71.4** | **105** | **100.0** |  |  |
| **Admission** |  |  |  |  |  |  | 5.559 | 0.018 |
| Isolation ward | 20 | 23.5 | 65 | 76.5 | 85 | 81.0 |  |  |
| Isolation ICU | 10 | 50.0 | 10 | 50.0 | 20 | 19.0 |  |  |
| **Total** | **30** | **28.6** | **75** | **71.4** | **105** | **100.0** |  |  |

\*Likelihood ratio

### Comorbid conditions of COVID-19 patients with regards to mortality

The comorbidities of COVID-19 patients were determined, the reported comorbid conditions were diabetes mellitus (DM), hypertension (HTN), DM with HTN and ischemic heart disease (IHD). There was no statistically significant association between comorbidities and COVID-19 mortality (p= 0.386, 0.083, 0.073, 0.872 respectively). Figure 1 below illustrates the percentage of deceased patients among each comorbid condition.

**Figuer 1:**
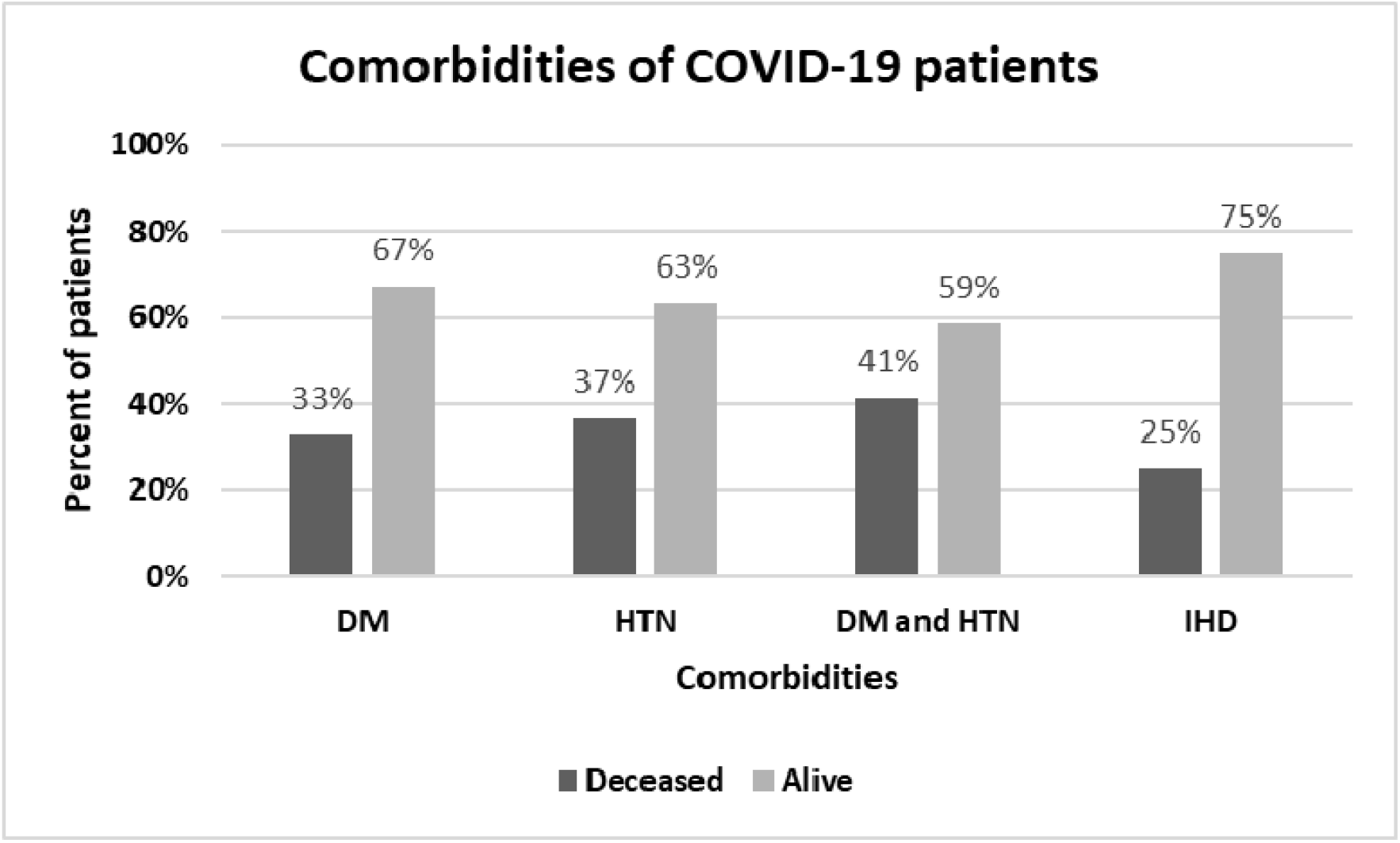
Comorbidities of COVID-19 patients based on the mortality status.

### COVID-19 presentation and treatment among participants based on their mortality status

COVID-19 presentation symptoms were assessed, they were divided into two categories. Typical symptoms, which were, fever, cough, fatigue and headache. Atypical symptoms, which were, gastrointestinal symptoms, reduced level of consciousness and neurological presentation. 81% (86/105) of the patients were presented with typical symptoms, while, 2.9% (3/105) of patients had atypical symptoms. In the other hand, 15.2% (16/105) of the patients had both typical and atypical symptoms. All (3/3) of the patients who had atypical COVID-19 presentation were deceased. Of the patients who had both typical and atypical symptoms, 31.3% (5/16) were deceased. Of the patients with regular symptoms, 25.6% (22/86) were deceased. A statistically significant association (likelihood ratio= 7.957, p=0.019) was found between COVID-19 presentation and mortality.

Association between the duration of hospitalization and complications was assessed with regards to mortality, but no statistically significant association was found (p> 0.05), table 2.

**Table 2:** Presentation and treatment of COVID-19 patients based on the mortality status (n=105)

| Variables | Mortality |  |  |  | Total | % | Likelihood ratio | p-value |
| --- | --- | --- | --- | --- | --- | --- | --- | --- |
|  | Deceased | % | Alive | % |  |  |  |  |
| **Presentation** |  |  |  |  |  |  | 7.957 | 0.019 |
| Typical symptoms | 22 | 25.6 | 64 | 74.4 | 86 | 81.9 |  |  |
| Atypical symptoms | 3 | 100.0 | 0 | 0.0 | 3 | 2.9 |  |  |
| Both | 5 | 31.3 | 11 | 68.8 | 16 | 15.2 |  |  |
| Total | 30 | 28.6 | 75 | 71.4 | 105 | 100.0 |  |  |
| **Duration of hospitalization** |  |  |  |  |  |  | 0.071* | 0.79 |
| <= 5 days | 20 | 27.8 | 52 | 72.2 | 72 | 68.6 |  |  |
| > 5 days | 10 | 30.3 | 23 | 69.7 | 33 | 31.4 |  |  |
| Total | 30 | 28.6 | 75 | 71.4 | 105 | 100.0 |  |  |
| **Complications** |  |  |  |  |  |  | 3.01 | 0.222 |
| Stroke | 1 | 20.0 | 4 | 80.0 | 5 | 4.8 |  |  |
| AKI | 0 | 0.0 | 4 | 100.0 | 4 | 3.8 |  |  |
| None | 29 | 30.2 | 67 | 69.8 | 96 | 91.4 |  |  |
| Total | 30 | 28.6 | 75 | 71.4 | 105 | 100.0 |  |  |
| **Enoxaparin dose** |  |  |  |  |  |  |  |  |
| **Therapeutic** |  |  |  |  |  |  | 0.031 | 0.859 |
| Yes | 4 | 26.7 | 11 | 73.3 | 15 | 14.3 |  |  |
| No | 26 | 28.9 | 64 | 71.1 | 90 | 85.7 |  |  |
| Total | 30 | 28.6 | 75 | 71.4 | 105 | 100.0 |  |  |
| **High prophylactic** |  |  |  |  |  |  | 9.528* | 0.002 |
| Yes | 11 | 17.5 | 52 | 82.5 | 63 | 60.0 |  |  |
| No | 19 | 45.2 | 23 | 54.8 | 42 | 40.0 |  |  |
| Total | 30 | 28.6 | 75 | 71.4 | 105 | 100.0 |  |  |
| **Prophylactic** |  |  |  |  |  |  | 9.774* | 0.002 |
| Yes | 11 | 57.9 | 8 | 42.1 | 19 | 18.1 |  |  |
| No | 19 | 22.1 | 67 | 77.9 | 86 | 81.9 |  |  |
| Total | 30 | 28.6 | 75 | 71.4 | 105 | 100.0 |  |  |
| **No prophylaxis** |  |  |  |  |  |  | 1.714 | 0.183 |
| Yes | 4 | 50.0 | 4 | 50.0 | 8 | 7.6 |  |  |
| No | 26 | 26.8 | 71 | 73.2 | 97 | 92.4 |  |  |
| Total | 30 | 28.6 | 75 | 71.4 | 105 | 100.0 |  |  |

\*Chi-square test

Enoxaparin doses used in the treatment of COVID-19 patients were evaluated. 14.3% (15/105) of the patients received therapeutic doses, 26.7% (4/11) of them were deceased. No statistically significant difference was found between mortality rate and enoxaparin therapeutic dose (*p*=0.859). 60% (63/105) of the patients received high prophylactic doses, 40 mg twice daily. Of those, 17.5% (11/63) were deceased, while 45.2% of the patients who did not receive high prophylactic doses of enoxaparin were deceased. A statistically significant difference was found between mortality rate and enoxaparin high prophylactic doses (*p*=0.002). Patients who received standard prophylactic enoxaparin doses were 18.1% (19/105), of those patients 57.9% (11/19) were deceased, while, 22.1% of these patients who did not receive prophylactic enoxaparin doses were deceased (*p*=0.002). In the other hand, 50% of patients who did not receive any anticoagulant were deceased (*p*= 0.183). Table 2 below, illustrates the association between COVID-19 presentation and treatment with regards to mortality.

### Odds ratio of the associated variables to COVID-19 mortality

All the variables associated with COVID-19 mortality were assessed, their odds ratios were described in table 3 below. Age, gender and comorbidities were assessed besides the admission and duration of hospitalization. As well as, the symptoms and complications of COVID-19 infection (table 3). Forest plot for the odds ratios of all the variables and their 95% confidence intervals was presented below (figure 2).

**Table 3:** Odds ratio of the associated variables to COVID-19 mortality.

| Variables | Mortality |  |  |  | Total | Odds Ratio (95% CI) |
| --- | --- | --- | --- | --- | --- | --- |
|  | Deceased | % | Alive | % |  |  |
| **Age:** |  |  |  |  |  | 0.268 ( 0.074-0.975) |
| ≤ 60 years | 3 | 12.0 | 22 | 88.0 | 25.0 |  |
| > 60 years | 27 | 33.8 | 53 | 66.3 | 80.0 |  |
| **Gender:** |  |  |  |  |  | 1.261 (0.518- 3.071) |
| Male | 20 | 30.3 | 46 | 69.7 | 66.0 |  |
| Female | 10 | 25.6 | 29 | 74.4 | 39.0 |  |
| **Admission:** |  |  |  |  |  | 0.308 (0.112- 0.845) |
| Isolation ward | 20 | 23.5 | 65 | 76.5 | 85.0 |  |
| Isolation ICU | 10 | 50.0 | 10 | 50.0 | 20.0 |  |
| **Duration of hospitalization:** |  |  |  |  |  | 0.885 (0.358- 2.184) |
| ≤ 5 days | 20 | 27.8 | 52 | 72.2 | 72.0 |  |
| > 5 days | 10 | 30.3 | 23 | 69.7 | 33.0 |  |
| **Comorbidities:** |  |  |  |  |  |  |
| **DM** |  |  |  |  |  | 1.455 (0.622- 3.403) |
| Yes | 16 | 32.7 | 33 | 67.3 | 49.0 |  |
| No | 14 | 25.0 | 42 | 75.0 | 56.0 |  |
| **HTN** |  |  |  |  |  | 2.129 (0.898- 5.046) |
| Yes | 18 | 36.7 | 31 | 63.3 | 49.0 |  |
| No | 12 | 21.4 | 44 | 78.6 | 56.0 |  |
| **DM+HTN** |  |  |  |  |  | 2.275 (0.917- 5.643) |
| Yes | 12 | 41.4 | 17 | 58.6 | 29.0 |  |
| No | 18 | 23.7 | 58 | 76.3 | 76.0 |  |
| **IHD** |  |  |  |  |  | 0.828 (0.083- 8.286) |
| Yes | 1 | 25.0 | 3 | 75.0 | 4.0 |  |
| No | 29 | 28.7 | 72 | 71.3 | 101.0 |  |
| **Complications:** |  |  |  |  |  | 1.423 (1.253- 1.615) |
| **AKI** |  |  |  |  |  |  |
| Yes | 0 | 0.0 | 4 | 100.0 | 4.0 |  |
| No | 30 | 29.7 | 71 | 70.3 | 101.0 |  |
| **Ischemic stroke** |  |  |  |  |  | 0.612 (0.066- 5.712) |
| Yes | 1 | 20.0 | 4 | 80.0 | 5.0 |  |
| No | 29 | 29.0 | 71 | 71.0 | 100.0 |  |
| **Symptoms:** |  |  |  |  |  |  |
| **Fever** |  |  |  |  |  | 0.459 (0.176- 1.194) |
| Yes | 20 | 24.7 | 61 | 75.3 | 81.0 |  |
| No | 10 | 41.7 | 14 | 58.3 | 24.0 |  |
| **Fatigue** |  |  |  |  |  | 1.130 (0.458- 2.791) |
| Yes | 10 | 30.3 | 23 | 69.7 | 33.0 |  |
| No | 20 | 27.8 | 52 | 72.2 | 72.0 |  |
| **Shortness of breath** |  |  |  |  |  | 0.717 (0.293-1.752) |
| Yes | 19 | 26.4 | 53 | 73.6 | 72.0 |  |
| No | 11 | 33.3 | 22 | 66.7 | 33.0 |  |
| **Headache** |  |  |  |  |  | 1.000 (0.183- 5.460) |
| Yes | 2 | 28.6 | 5 | 71.4 | 7.0 |  |
| No | 28 | 28.6 | 70 | 71.4 | 98.0 |  |
| **Cough** |  |  |  |  |  | 0.509 (0.208-1.248) |
| Yes | 18 | 24.3 | 56 | 75.7 | 74.0 |  |
| No | 12 | 38.7 | 19 | 61.3 | 31.0 |  |
| **Reduced consciousness** |  |  |  |  |  | 2.607 (0.350- 19.415) |
| Yes | 2 | 50.0 | 2 | 50.0 | 4.0 |  |
| No | 28 | 27.7 | 73 | 72.3 | 101.0 |  |
| **GI symptoms** |  |  |  |  |  | 1.833 (0.590- 5.697) |
| Yes | 6 | 40.0 | 9 | 60.0 | 15.0 |  |
| No | 24 | 26.7 | 66 | 73.3 | 90.0 |  |

**Figuer 2:**
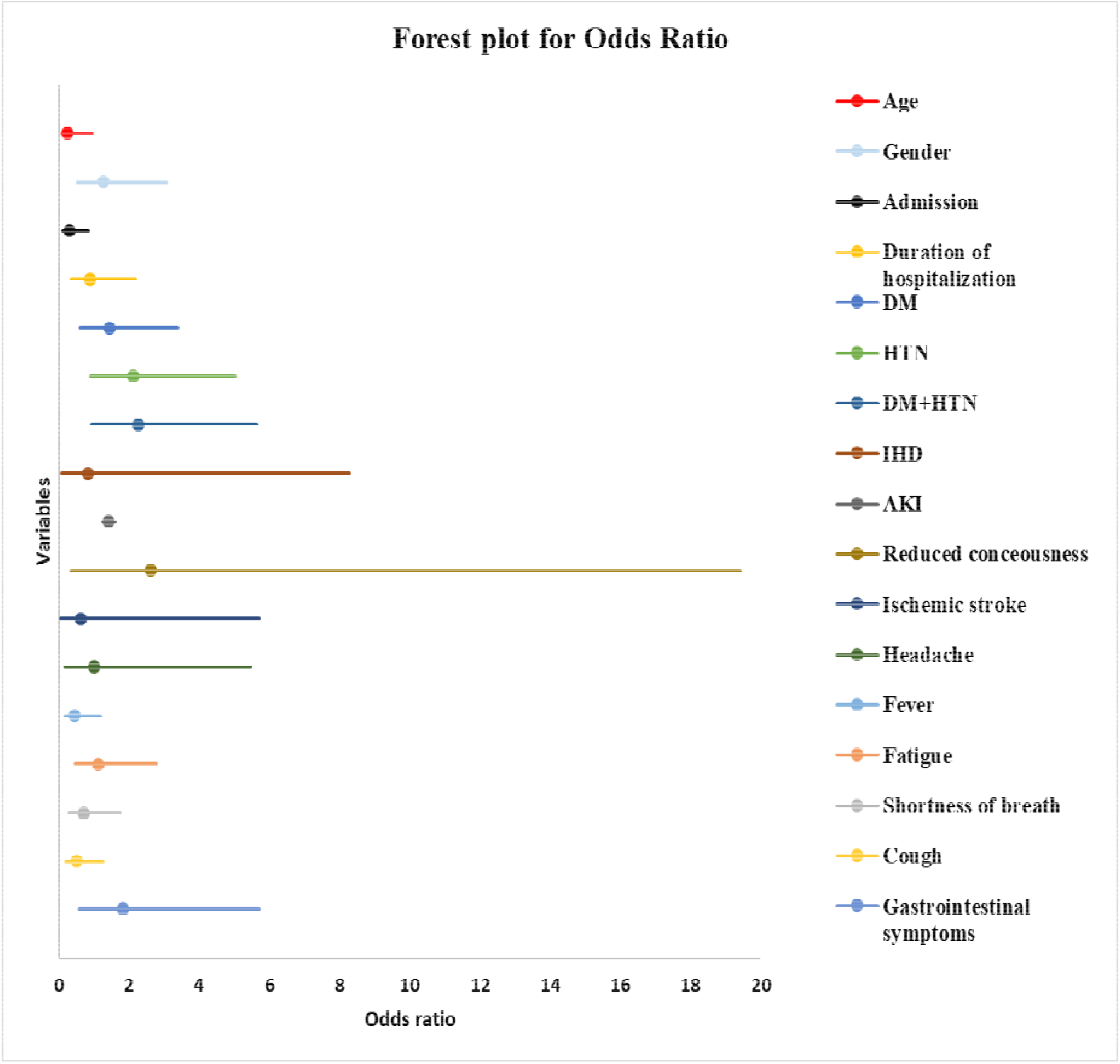
Forest plot for odds ratio of variables associated to COVID-19 mortality.

### Logistic regression model for predicting the mortality among COVID-19 patients

Our equation for the regression model fitted the model perfectly (Chi^2^= 28.030, *p*= 0.001), the model equation was as follows:

Mortality = −14.724+ (1.387* Age) + (−0.323* Gender) + (1.814* Admission) + (0.193* Ischemic Heart Disease) + (−0.369* Fever) + (1.595* Cough) + (1.953* Complications) + (0.149* Duration of hospitalization) + (0.999* Enoxaparin dose).

Age, admission ward, cough and enoxaparin dose were statistically significant predictors for COVID-19 mortality (*p*= 0.014, 0.011, 0.015, 0.006 respectively). In the other hand, gender and fever had negative influence to mortality. Although, complications was not a statistically significant factor (*p*=0.100), it contributed by more than seven times in our model (OR=7.051, CI: 0.69-72.046). Table 4 below illustrates the predicting factors of the logistic regression model for COVID-19 mortality.

**Table 4:**
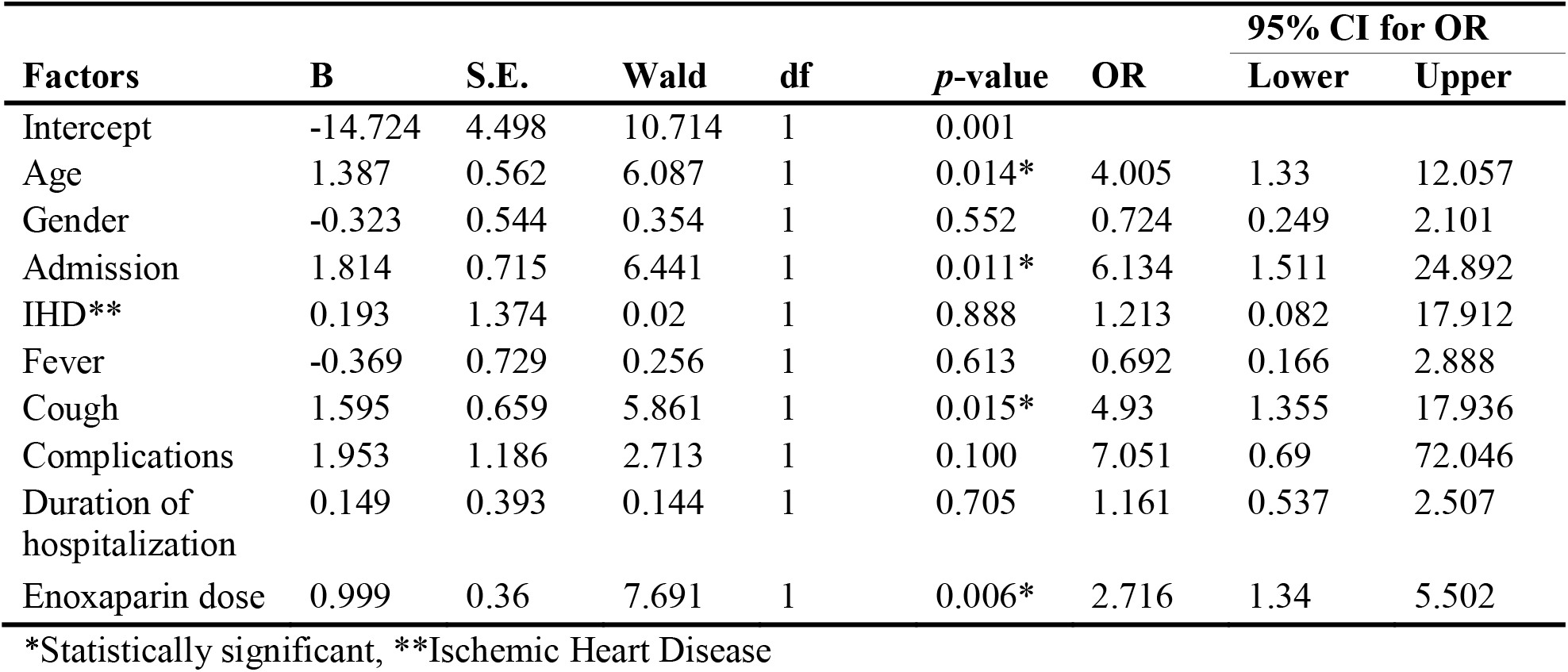
Mortality prediction model for COVID-19 patients.

## Discussion

Mortality rate among Sudanese patients with COVID-19 infection was (29%), this was higher than reported rates (17.6% and 3.9%) in published studies [8, 12]. However, this rate was lower than the rate reported in Brazil [19].

Age was significantly associated with mortality (*p*=0.034), consistent with [1, 20]. None of the participants who aged ≤ 45 years had died, consistently with a developed mortality score [16] in which, patients aged below 50 years had no points in the score. While, 33.3% of patients above 60 years were deceased (p= 0.034). Our results were comparable to the reported mortality rates (11.7% and 54.2%) for age groups between 65-69 years and above 80 years, respectively [6]. In a Spanish study [4] assessing the association between age and mortality, 95.3% of deceased COVID-19 patients aged 60 years and above, (*p*=0.006).

Gender had no statistically significant association with mortality, this was in contrast with published studies [1, 16, 20] and in line with an English and a Brazilian studies, in which, mortality rates were the same among males and females [19, 21].

In our study, mortality among severe cases (19%) was higher than the rates reported in a Japanese study (12%) and in a Vietnamese study (16%) among severe cases [22, 23]. 19% of our patients were admitted to the isolation ICU, this percent was higher than the percent reported (12%) in a cohort study [12] and similar to another study [13], in which, critical cases were (19.5%) of patients. Patients who were admitted to the ICU had higher mortality rate (50%) than patients who were admitted to the general ward (23.5%).

The reported comorbidities were, DM, HTN, DM+HTN, and IHD. These were similar to reported comorbidities [8, 24] besides other comorbidities. In our study, (33%) of the diabetic patients were deceased, while, in Brazil, the rates were higher [19]. The odds ratio for diabetes as a related factor to mortality among our patients was 1.455 (95% CI=0.622-3.403, p= 0.386), while in another study [2], it was 0.099 (95%CI=0.016– 0.627, *p*=0.014). Interestingly, when we assessed the association between comorbidities of the patients and COVID-19 mortality, no statistically significant association was found between these comorbidities and mortality (*p*> 0.05). Consistent with the results reported [1, 4, 12]. Contrary to a study reported that DM and HTN had a statistically significant association with mortality [20]. In our study, the percent of COVID-19 patients with hypertension who were deceased was 37%, this was comparable to the percentages reported in a systematic review [11], in which, COVID-19 mortality rate in patients with HTN ranged between (30.5%) and (64.7%).

Symptoms of COVID-19 presentation were assessed in our patients. The patients who presented with atypical symptoms, as gastro-intestinal (GI) symptoms and reduced consciousness, had the highest mortality rate (100%). GI symptoms had an odds ratio of 1.833 (95% CI=0.590-5.697).While, in a published study [25], the odds ratios for nausea and vomiting were (0.63 [0.38-1.04] and 0.92 [0.59-1.44] respectively). Patients who were presented with typical symptoms had (25.6%) mortality rate.

The duration of hospitalization had no statistically significant association with mortality. As well as, the complications, as stroke and acute kidney injury (AKI), caused by COVID-19 infection had no statistically significant association to mortality. As in a study, (15%) of patients who died from COVID-19 infection had no complications [4].

Standard thrombo-prophylaxis is not enough for COVID-19 patients [26]. Patients in our study, who had no anticoagulation treatment had (50%) mortality rate (*p*=0.183), while patients who received high prophylactic doses of enoxaparin, 40 mg twice daily, had the lowest mortality rate (17.5%; *p*= 0.002). What interesting is, the mortality rate of patients who received therapeutic enoxaparin doses was higher than in patients who received high prophylactic doses. This was consistent with the results of a published study in the United States [27], in which, prophylactic enoxaparin dose had statistically significant difference in mortality (*p* =0.001), while therapeutic dose had no statistically significant difference (*p*=0.57). Clinical trials are still being conducted to compare the efficacy between prophylactic and therapeutic doses of enoxaparin among COVID-19 patients [28, 29].

The prediction of mortality among our patients was performed through logistic regression. Our model had perfect fit (*p*= 0.001). Age was a statistically significant factor, consistent with published literature [7, 12, 24, 30]. Admission, cough and enoxaparin dose were statistically significant prediction factors (*p*< 0.05). Interestingly, gender was not a significant predictor of mortality in our study patients, this was contrary to the results of published prediction models [7, 12]. Although, complications was not statistically significant (*p*=0.100), it contributed by more than seven times in our model (OR=7.051, CI: 0.69-72.046). Fever was not statistically significant, in line with a published model [12] and in contrast with [7]. In our model, IHD was not a statistically significant predictor contrary to a published study [24].

The limitations of our study were the size of the study, as it was a single centre study. The data collection sheet was not standardized, only validated through an expert in research methods. Besides the retrospective nature of data collection which might affect the quality of the data collection.

## Conclusions

Age, admission, cough and enoxaparin dose were statistically significant prediction factors (*p*< 0.05) for mortality rate among hospitalized COVID-19 patients. Interestingly, gender was not a significant predictor of mortality in our study patients, alongside with comorbidities.

## Recommendations

Special care must be provided for patients older than 60 years and intensifying the preventive measures for this slide of the community as they are the vulnerable population group. Multi-centre wider studies to confirm these results will be of high significance. Also, assessment of enoxaparin prophylactic and therapeutic doses in a direct cause-effect study must be conducted to define the most suitable dose for hospitalized COVID-19 patients.

## Data Availability

All supporting data are available.

## Abbreviations

AKI: Acute Kidney Injury
DM: Diabetes Mellitus
GI: Gastro-Intestinal
HTN: Hypertension
ICU: Intensive Care Unit
MOH: Ministry Of Health
IHD: Ischemic Heart Disease
IRB: Institutional Review Board
OCHA: Office for the Coordination of Humanitarian Affairs

## Declarations

### Ethics Approval and Consent to Participate

Ethical approval was obtained from the Medical administration of Imperial Hospital. The study proposal was submitted to the research administration at the State Ministry Of Health (MOH), Khartoum. Expedited review was conducted by the IRB of the MOH and approval was granted. Another copy of the proposal was submitted to the Administration of Private Medical Facilities. Regarding participants, the data were collected retrospectively from medical records and their confidentiality was assured with the use of an anonymous research tool. Informed consents from the patients/ surrogate decision makers were obtained by contacting them through their registered phone numbers. The collected data were used strictly for the purpose of the study objectives.

### Consent for publication

All authors have read the final manuscript and gave their consent for the article to be published in Infection. No clinical details of participants that might compromise their anonymity were used in the development of this manuscript titled **“Prediction of COVID-19 mortality among hospitalized patients in Sudan”**.

### Availability of supporting data

All supporting data are available.

### Authors’ contributions

All authors have read the final manuscript and gave their approval for publication.

**GOHA**: Wrote the manuscript, conducted the analysis and assisted in the data collection.

**MAAY**: Collected the data, assisted in getting the ethical approval, read the first draft of the manuscript and proof read the final manuscript prior to submission.

**DSIM**: Facilitated the administrative arrangements at Imperial Hospital, assisted in the data collection and read the final manuscript prior to submission.

## Acknowledgments

The authors are grateful to Shurouq O. H. Abdelraheem, Faculty of Engineering, University of Khartoum for her assistance in the data entry process. The authors also acknowledge the cooperation of the departments of Imperial Hospital during the data collection.

